# Quantitative multimodal microstructural imaging associations with chronic stroke motor impairment

**DOI:** 10.1101/2025.05.22.25326475

**Authors:** Zeena-Britt Sanders, Melanie K Fleming, Tom Smejka, Marilien C Marzolla, Daniel Papp, Alberto Lazari, Anderson M Winkler, Emily Moore, Martina F. Callaghan, Heidi Johansen-Berg, Cassandra Sampaio Baptista

## Abstract

**Background:** Corticospinal tract (CST) integrity, assessed using diffusion weighted MR imaging, has frequently been associated with motor impairment following stroke. However, which specific biological features contribute to this relationship remains unknown. In the current paper we used quantitative multimodal imaging to shed light on the biophysical properties underlying this relationship in the CST and explore additional whole brain relationships with motor impairment.

**Methods:** Twenty-seven chronic stroke survivors underwent MRI scanning, including multi-shell diffusion weighted imaging and quantitative Multi-Parameter Mapping, from which we derived ten metrics, sensitive to different microstructure properties. We tested the relationship between these MRI metrics and stroke survivors’ impairment scores on the Upper-Extremity Fugl Meyer (UE-FM), using multimodal and unimodal statistical inference, in both the CST and across the whole brain.

**Results:** Using an ROI approach to replicate and extend on previous findings, a joint multimodal analysis revealed that greater CST microstructure asymmetry was associated with worse motor impairment. The strongest relationships were found between impairment and several modalities sensitive to free water and myelination. This result was further confirmed using a voxel-wise approach. Beyond the CST, modalities sensitive to free water, myelination and the Neurite Density Index, were also found to be related to motor impairment, particularly in the superior longitudinal fasciculus, corpus callosum and thalamic radiations, with magnetisation transfer saturation showing the strongest relationship in these areas.

**Conclusion:** Modalities that are sensitive to myelin and free water exhibited the most significant correlation with motor impairment. This suggests that these specific biological features may account for the previously identified relationships between diffusion metrics and motor impairments. Myelin metrics in particular were also highly correlated with motor impairment in other brain regions. This study illustrates the advantages of using multimodal data to identify more specific biological factors that contribute to motor impairment in stroke patients.

## Intro

Stroke is a leading cause of disability worldwide^1^, often resulting in motor impairment that affects the quality of life of survivors^2^. Understanding the underlying mechanisms of motor impairment and identifying potential biomarkers for personalized treatment and recovery are essential for improving rehabilitation strategies.

One of the key findings in stroke neuroimaging research is the relationship between corticospinal tract (CST) integrity and motor impairment^3–6^. For example, the asymmetry in fractional anisotropy (FA) of the CSTs has been found to be a significant predictor of functional recovery potential in chronic stroke survivors, with no meaningful gains possible if FA asymmetry exceeds a value of 0.251^7^ (see also^8^). The use of such a region of interest (ROI)-based asymmetry approach has the benefit of being able to deal with anatomical variation across individuals. Such insights have led to the proposal of CST integrity as a biomarker for personalizing treatments and assessing recovery potential^9^.

One of the challenges in this field is that diffusion weighted imaging (DWI) is sensitive to multiple underlying biological mechanisms^10,11^, making it difficult to disentangle the specific effects that contribute to motor impairment. Previous research has linked several biological mechanisms to motor impairment and recovery after stroke including myelination^12–14^, axonal remodelling^15,16^ and iron accumulation^17,18^. Multimodal imaging approaches can shed light on underlying biological mechanisms by including modalities with differing sensitivity to biological tissue properties^19^.

In the current paper we used a multimodal imaging approach to elucidate the tissue properties contributing to the commonly reported relationship between CST integrity and motor impairment after stroke. We specifically acquired multi-shell DWI and derived Diffusion Tensor Imaging (DTI) and Neurite Orientation Dispersion and Density Imaging (NODDI) metrics^20^. DWI measures the directional dependence of water diffusion and can be used to calculate maps of fractional anisotropy (FA), mean diffusivity (MD), axial diffusivity (AD) and radial diffusivity (RD). FA measures diffusion anisotropy which reflects the amount of diffusion occurring in the principal diffusion direction compared to the secondary and tertiary direction. FA is thought to be sensitive to axonal integrity, axon diameter and density and myelination^10,11^. MD measures the average amount of diffusion across all three diffusion directions and is sensitive to edema, cellularity and necrosis^21^. AD reflects the amount of diffusion occurring in the principal diffusion direction and is proposed to be sensitive to axonal degeneration, whereas RD measures diffusion occurring perpendicular to the principle diffusion direction and is thought to be sensitive to degeneration of myelin^22,23^. Using the NODDI model we derived maps of Neurite Density Index (NDI) which reflects the packing density of axons and dendrites, Orientation Dispersion Index (ODI) reflecting the orientational coherence of the neurites, and isotropic fraction (FISO; free water volume fraction) reflecting cerebrospinal fluid (CSF)^20^.

In addition, we acquired Multi-Parameter Mapping (MPM)^24^. We used the hMRI toolbox^25^ to extract quantitative maps of Magnetization transfer saturation (MTsat) which has been shown to be strongly related to myelination^26^, R1 (=1/T1, where T1 is the longitudinal relaxation time) which is sensitive to myelin, iron and water content, and R2* (=1/T2*, where T2* is the effective transverse relaxation time) which is considered to be a marker for iron^24,25^.

Using this data, we carried out unimodal and multimodal analyses to address the following aims:

1. Confirm the association between asymmetry of the CST microstructure (ROI analysis) and motor impairment.
2. Test for an association between brain-wide white matter asymmetry (voxelwise analysis) and motor impairment.
3. Test for an association between brain-wide white matter structure (voxelwise analysis) and motor impairment.

## Methods

### Participants

Twenty-seven stroke survivors (> 6 months post stroke) with mild-moderate upper limb motor impairment were recruited to take part in a clinical trial^27^ (ClinicalTrials.gov: NCT0377515) investigating the effect of functional MRI neurofeedback on motor performance and impairment. Results from the trial and intervention are reported separately^27^ and are not a focus of the current paper. For the current paper, only the DWI, the MPM and the Upper-Extremity Fugl-Meyer (UE-FM) data from these participants were considered to separately investigate how the multimodal MRI data relates to motor impairment in stroke patients.

The data was collected both at the baseline session, one week before the intervention, and the follow-up session, one week after the last day of intervention. There was approximately three weeks between the baseline and the follow-up session. The sessions were identical in terms of measures acquired and the order of events. As the MPM sequences were sensitive to pulsatile motion, we selected the best quality MRI data (MPM and DWI) from the session (either baseline or follow up) with the highest average MPM quality (see MRI-preprocessing below for more details).

Three participants were excluded from the study (1 participant did not complete the full scan due to discomfort, 2 participants had low data quality in both sessions due to motion artefacts), resulting in 24 participants being included (19 males) in the current study. Demographic information for these participants is presented in **Table 1**.

**Table 1:** Demographic information for participants. Note: maximum score on the UE-FM is 66 and lower scores are indicative of greater impairment of the affected upper limb.

|  |  |
| --- | --- |
| Mean age, years (range) | 60.2 (36-86) |
| Male:female | 19:5 |
| Mean months post-stroke (range) | 74.8 (7-237) |
| Hand affected | R=8, L=16 |
| Multiple strokes | Y=2, N=22 |
| Mean of two sessions of UE-FM (range) | 45.07 (21.00-62.75) |

### UE-FM

The UE-FM was scored by two independent blinded assessors and the average score was taken for each session. As there was no evidence of any change on this measure over the trial, the average score between the two sessions was calculated for the current analysis. Lower scores on the UE-FM are indicative of greater motor impairment of the affected arm (max score 66). Note: in the clinical trial no significant interaction effects between timepoint and intervention type were detected for the UE-FM score. A main effect of time fell short of significance (p =0.051). For more details, see the mean and standard error for timepoints and groups in Sanders et al., 2022 supplementary information^27^.

### MRI data acquisition

All MRI data were acquired on a 3T Siemens Magnetom Prisma scanner (Siemens AG) using a 32-channel head coil at the Oxford Centre for Functional MRI of the brain (FMRIB). DWI data was acquired using a multi-shell 2D EPI sequence with both A>P and P>A phase encoding in two separate runs (FOV=214×214mm^3^, 76 slices of 1.75mm, TR=2483ms, TE=78.2ms, voxel-size=1.75×1.75×1.75 mm^3^, multiband factor=4, 11 *b*∼0s/mm volumes, 60 different diffusion directions for b-value=1500 s/mm and b-value=2500 s/mm each interspersed per run: 120 directions total).

The multi-echo 3D MPM protocol included the acquisition of proton-density weighted (PDw), MT-weighted (MTw) and T1-weighted (T1w) images (FOV= 256×224×180 mm^3^, TR=25ms, first TE=2.3ms, echo spacing=2.3ms, number of echoes=8,6,8 for PDw, MTw and T1w respectively, voxel-size=1.5×1.5×1.5 mm^3^, flip angle=6° for PDw and MTw, flip angle=21° for T1w, GRAPPA acceleration factor 2×2, with 40 reference lines in each direction). MTw scans were preceded by an off-resonance Gaussian MT pulse with a 2kHz offset, 4ms duration and 220° flip angle. After the first five participants had been scanned, we additionally acquired receive-coil sensitivity field maps from the body and head coil in order to correct for inter scan motion^28^. This was reflected in the later group analysis by including a covariate (see below).

A T1w 3D MPRAGE image (henceforth: T1; FOV 192×174×192 mm^3^, TR=1900ms, TE=3.97ms voxel-size=1×1×1mm^3^) was acquired and used for registration purposes and to manually delineate the lesioned tissue^27^.

### MRI pre-processing

DWI data were pre-processed as in Sanders et al., (2022)^27^ using FMRIB’s Diffusion Toolbox (FDT). Distortion correction was carried out using *topup* and eddy-current correction was applied using *eddy*. *Dtifit* was used to fit a simplified diffusion kurtosis model at each voxel which resulted in whole brain maps of FA, MD, AD and RD. Additionally, the NODDI model^20^ was estimated using FSL’s CUDIMOT based on the Bingham-NODDI distribution^29^.This resulted in maps of NDI, ODI and FISO. MPM data was preprocessed using the hMRI toolbox^25^ implemented in SPM (version 12). This resulted in whole brain maps of magnetization transfer saturation (MTsat), R1 and R2*. The hMRI quality assessment tool was used to assess the quality of the maps produced and to decide which dataset to use (baseline or follow-up session). In particular, we considered the standard deviation in the white matter of the R2* map (SD-R2*), which is sensitive to intra-scan motion^30^, and selected the dataset with the lowest standard deviation. The average SD-R2* of the selected datasets was 4.47, compared to 4.77 of the non-selected datasets with average SD-R2* values (over the multi-echo acquisitions) ranging from 3.68 to 6.34.

### ROI registration

To assess how CST asymmetry is related to scores on the UE-FM, CST regions of interest (ROIs) were delineated in MNI standard space from the JHU white-matter tractography atlas (see inset Figure 1). These ROIs were then registered into native participant space using the steps described below.

**Figure 1:**
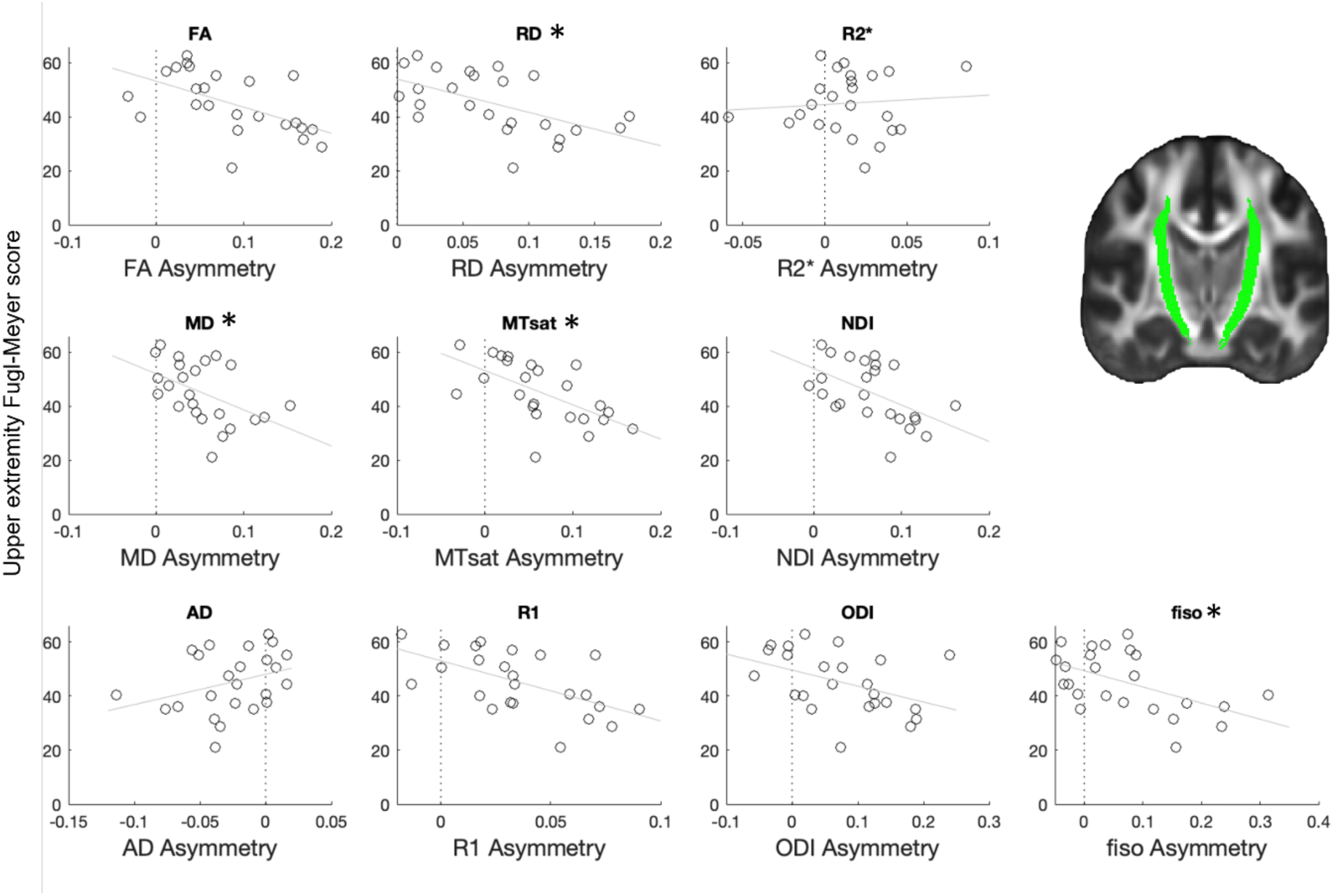
Scatter plots (with least squares fit line) of the CST asymmetry value and UE-FM score for each modality. † Note that signs of MD, RD, ODI and FISO are negated. CST masks (green) were used to calculate asymmetry values. A joint analysis using NPC reveled a significant multimodal association between CST asymmetry and with UE-FM (p=0.0032, corrected). Unimodal relationships that survived multiple testing correction across modalities (i.e. RD, MD, MTsat and FISO) are signified with an asterisk.

Registration for ROI analysis was performed as in^27^. Briefly, the FA maps for each day were linearly registered to the structural MPRAGE T1 image of the same day using FSL’s FLIRT. T1 images from both days were then transformed into a halfway midspace between the two days and averaged to create a ‘midstruct’ image. This image was created by first calculating the halfway linear registration between the T1 images from the baseline and follow-up sessions. The T1 images were then linearly registered to the halfway space and averaged to create a participant-specific midstruct image. These linear transformation steps were concatenated and inverted. The midstruct image was then non-linearly aligned to the MNI standard space using ANTs (Advanced Normalization Tools; ^31^) as this tool produced superior registrations than FNIRT due to the presence of the stroke lesions. Registration via the midstruct image was used to avoid bias in the longitudinal analysis carried out in the original clinical trial. The same registrations were kept in the current paper. The inverse transformations were then used to extract the CST ROIs into participant and day specific native space. For the MPM data an additional registration step was carried out between the MTsat map and the FA map, which was then used to linearly register the MPM data to the FA map using FLIRT.

If a lesion encroached on the CST ROIs, the overlapping voxels were removed from the ROI, and MRI metrics were extracted for each voxel within the remaining CST tract. Outlier values were removed (>3xIQR (interquartile range)) and the mean values were calculated for the affected (CST_aff_) and the unaffected (CST_unaff_) tract for each modality. CST asymmetry was calculated for each modality using these mean values (CST_unaff_ – CST_aff_)/ (CST_unaff_ + CST_aff_).

### Whole brain registrations

Registration for voxel-wise analysis was performed as in^27^. First, data was flipped to align all the affected hemispheres. The FA maps were then linearly registered to the halfway space between the baseline and follow-up sessions and averaged to create a participant-specific midspace FA image. For each participant, the midspace image was non-linearly registered to the FMRIB FA template using ANTs. Lesion masks were used to exclude lesion areas from the registration. TBSS^32^ was used to extract the white matter skeleton for each participant. For all voxelwise analyses, areas with high lesion overlap across participants were removed. This was done by registering all participants’ lesion maps to the FA template, averaging the lesion maps, and removing areas from the skeleton mask where more than one third of participants had lesions. The registration was applied to the other DTI modalities (including NODDI maps) and MPM data using the non-FA standard TBSS approach^32^. To perform an asymmetry analysis, as implemented in TBSS^32^, a symmetrical skeleton was created, and data was projected onto this skeleton before the left minus right subtraction occurred. This resulted in skeletonised data for half of the brain which reflected the differences in values between the left and right hemisphere (similar to the CST asymmetry used in the ROI analysis). Note that data from right hand affected stroke survivors was flipped, therefore for all survivors this subtraction corresponded to the affected hemisphere minus the unaffected.

### Statistical Analysis with Permutation Analysis of Linear Models

For both ROI and voxelwise analyses, FSL’s Permutation Analysis of Linear Models (PALM^33^) was used to carry out joint inference using non-parametric combination (NPC) with Fisher’s combining function over the multiple modalities (multimodal inference) and to simultaneously perform unimodal tests with family-wise error rate correction (FWER) over multiple modalities. This is done within PALM by calculating the permutation distribution of the extremum statistics across all modalities and then producing adjusted p-values.

The structural metrics employed here model different biophysical properties, therefore different relationships with impairment for each modality are expected. For instance, we expect that low FA, MTsat, R1, R2*, NDI to be associated with high impairment but the opposite relationship for MD, RD, ODI and FISO. Whilst for all other modalities clear predictions could be made in terms of expected direction of effect, this was not the case for AD where both increased and decreased AD has been associated with axonal damage and neurodegeneration^22,23,34,35^. Therefore, in order to test for directionally specific joint effects in the NPC analysis as implemented by PALM^33^, we were required to negate MD, RD, ODI and FISO by multiplying each value by −1, so that in all cases a negative correlation could be hypothesised between the imaging derived markers and impairment. We did not negate AD as we cannot be sure of the direction of the prediction as explained above. This allows for contrasts to be set to test for *joint* associations in the hypothesised direction between the modalities and UE-FM, as well as in the opposite direction.

Our first aim was to confirm the association between asymmetry of the CST microstructure and motor impairment (two-session average UE-FM score) using an ROI approach. As MD, RD, ODI and FISO values were negated, all modalities mean values are expected to be lower in the stroke-affected side than the unaffected due to damage and degeneration to the stroke affected hemisphere. Therefore, positive asymmetry scores (e.g. lower FA in the affected side than the unaffected) are expected for these modalities and a negative correlation with impairment (UE-FM) scores (e.g. greater asymmetry related to lower scores on the UE-FM). For this ROI-based analysis 5000 permutations and a p-value of <0.05 were selected (FWER-corrected across modalities)^36^.

Two separate voxelwise analyses were carried out to address aims 2 and 3. One to investigate white matter asymmetries and the other on the whole-skeleton data. For the two voxel-wise analyses, NPC with Fisher’s combining function was used in PALM to carry out multi-modal (p<0.05 corrected across all voxels), and simultaneous unimodal tests (p<0.05 corrected across all modalities and voxels) on the association between all the MRI modalities and UE-FM score in the hypothesised direction as well as the opposite direction. Threshold-free cluster enhancement (TFCE) and 1000 permutations (lower number of permutations were set because the voxel-wise multimodal analysis was computational demanding) were used to determine FWER-corrected p-values. Additionally, the tails approximation option in PALM was used to accelerate permutation testing^37^.

### Covariates

For both the ROI analysis and the voxelwise analyses we included the following covariates. Age was included as a covariate as age has been shown to be associated with changes in white matter microstructure ^38,39^ which are detectable using the MPM sequence^40^. Since the time-since-stroke of our participants varied widely from less than a year to more than 15 years (Table 1), we also included the reciprocal of the time-since-stroke as a covariate under the assumption that there would be greater impairment closer to the time of stroke than later. Finally, as mentioned above, a covariate was included to reflect the addition of receive-coil sensitivity maps after the first five participants were scanned. Results without these three covariates can be found in supplementary materials.

## Results

### Multimodal signature in CST Asymmetry

We first tested relationships between CST asymmetry of the ten MRI modalities and UE-FM score, using PALM to examine multi- and uni-modal effects. We predicted that CST asymmetry measured using FA, AD, MTsat, R1, R2* and NDI would have a negative relationship with UE-FM score (higher asymmetry associated with lower scores on the UE-FM), whereas we predicted a positive relationship between CST asymmetry measured using MD, RD, ODI and FISO and UE-FM. We negated MD, RD, ODI and FISO in order to make joint inferences with NPC testing^36^.

The NPC analysis revealed a significant relationship (p=0.0032, corrected) between the multimodal MRI CST asymmetry metrics and the UE-FM, showing a common association across the modalities, such that greater asymmetry scores were associated with worse motor impairment. The unimodal analysis (**Table 2**) revealed significant relationships between MD, RD, MTsat and FISO asymmetry and UE-FM, after FWER correction for multiple testing across modalities (see **Table 2** for full statistics and **Supplementary Table 1** for results from the opposite contrast). Scatter plots for each modality and UE-FM score are shown in **Figure 1**.

**Table 2:** T-statistics and FWE corrected (across modalities) and uncorrected p-values for each modality in the ROI-based CST asymmetry analysis.

|  | t-stat | uncorrected p-value | corrected p-value |
| --- | --- | --- | --- |
| **FA** | 2.6872 | 0.0088* | 0.0554 |
| **MD** | 2.8445 | 0.0060* | 0.0408* |
| **AD** | -1.8454 | 0.9578 | 1.000 |
| **RD** | 3.2616 | 0.0038* | 0.0192* |
| **MTsat** | 3.0346 | 0.0042* | 0.0280* |
| **R1** | 2.6417 | 0.0100* | 0.0594 |
| **R2\*** | -0.3155 | 0.6154 | 0.9980 |
| **NDI** | 2.7493 | 0.0096* | 0.0482 |
| **ODI** | 1.7056 | 0.0548 | 0.2758 |
| **FISO** | 2.9087 | 0.0068* | 0.0358* |

### White matter asymmetry

PALM was used to carry out a voxelwise multi- and uni-modal analysis of relationships between white matter asymmetries across the whole brain and UE-FM scores. Asymmetry was assessed as the voxelwise difference between left and right hemispheres, which in this study corresponds to the affected hemisphere minus the unaffected.

No significant result was found when testing for a joint effect of the ten modalities using NPC (peak voxel p=0.057 in the superior longitudinal fasciculus (SLF), corrected, **Supplementary Figure 1**). When covariates were not included in the analysis this result reached significance in the CST, the SLF and in the superior thalamic radiation (see **Supplementary Figure 2**).

The unimodal analysis revealed a negative association, as predicted, for MD, RD, MTsat, R1 and NDI (p<0.05 corrected across all voxels, **Figure 2**; a trend was found in FA, with peak voxel p=0.051, **Supplementary Figure 3**), indicating that greater asymmetry is associated with lower scores on the UE-FM. This relationship was found across several tracts including the CST, the SLF, anterior and superior thalamic radiation and the frontal aslant tract (FAT; **Figure 2**). A positive association between UE-FM scores with AD was found to be significant in the SLF (p<0.05 FWE corrected across all voxels, **Figure 2**). For analysis without covariates see **Supplementary Figure 4**.

**Figure 2:**
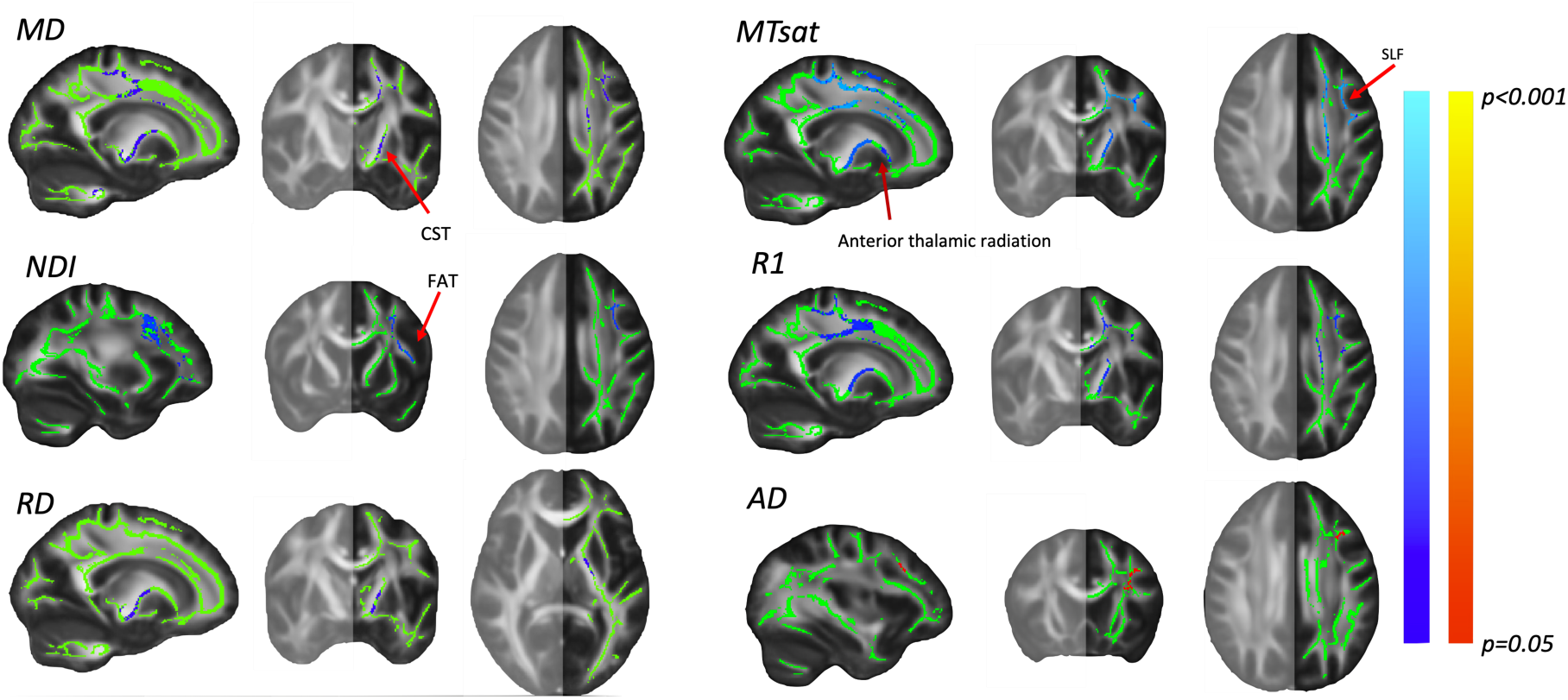
White matter asymmetry, unimodal analysis. A significant negative association (shown in blue) was found between MD, RD, MTsat, R1 and NDI asymmetry, and Fugl-Meyer upper extremity test (note that signs of MD, and RD values were negated). Significant voxels were found in several motor related areas including the CST, SLF, FAT and superior and anterior thalamic radiation. Additionally, a significant positive association (shown in red) was found between AD and UE-FM. Significant clusters are only projected in one hemisphere as asymmetry analysis considers the difference between affected hemisphere minus the unaffected.

After FWER across modalities only MTsat survived (**Figure 3**; for analysis without covariates see **Supplementary Figure 5**).

**Figure 3:**
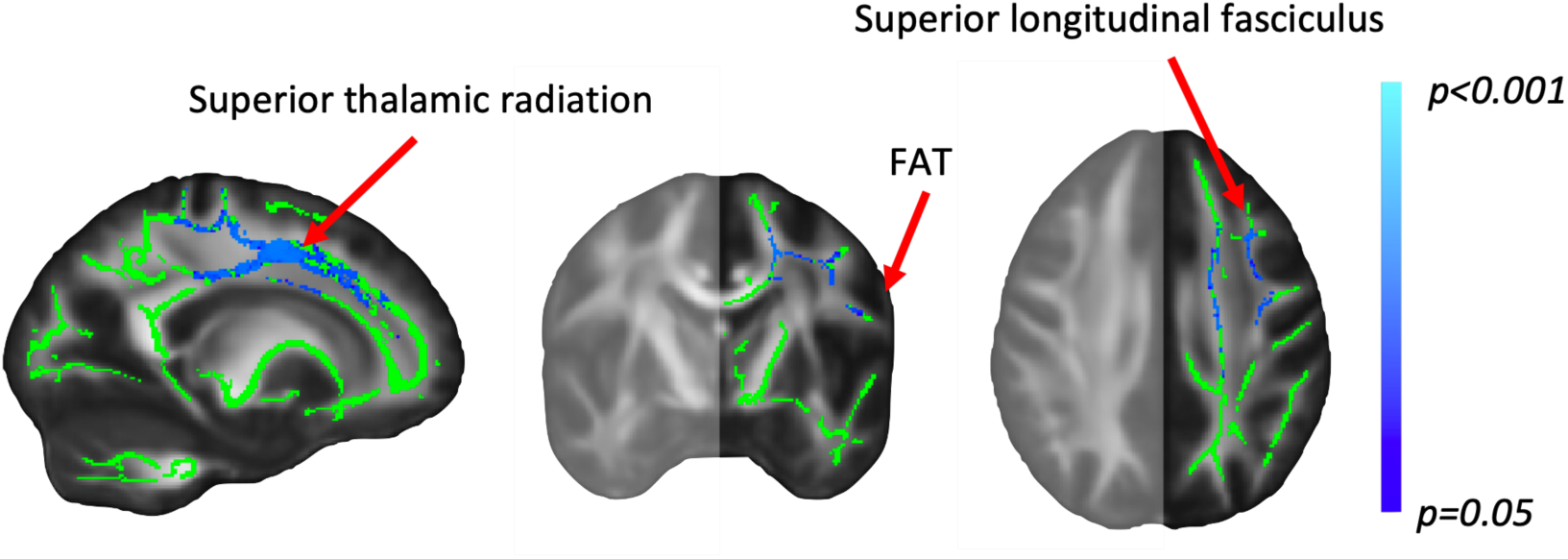
The negative association between MTsat asymmetry and UE-FM score survived multiple testing correction across modalities. Significant voxels were found in several motor related areas including the SLF, FAT and superior thalamic radiation. Significant clusters are only projected in one hemisphere as asymmetry analysis considers the difference between affected hemisphere minus the unaffected.

### Whole white matter analysis

Finally, we used a voxelwise NPC approach to test relationships between the whole white matter skeleton and UE-FM. No significant result was found when testing for a joint effect of the ten modalities using NPC (for analysis without covariates see **Supplementary Figure 6**).

The unimodal tests revealed a significant positive association between UE-FM and FA, MD, RD and NDI (**Figure 4**; p<0.05 corrected across all voxels; for analysis without covariates see **Supplementary Figure 7**) in several brain tracts in the stroke-affected side, indicating that higher values of FA and NDI, and lower values of MD and RD, are associated with less motor impairment. For FA, a significant positive association was found in the lower portion of the stroke-affected CST, whereas for the remaining modalities positive associations were also found in the superior portion of the CST, the corpus callosum and the SLF (**Figure 3**). None of these results survived FWER correction across modalities.

**Figure 4:**
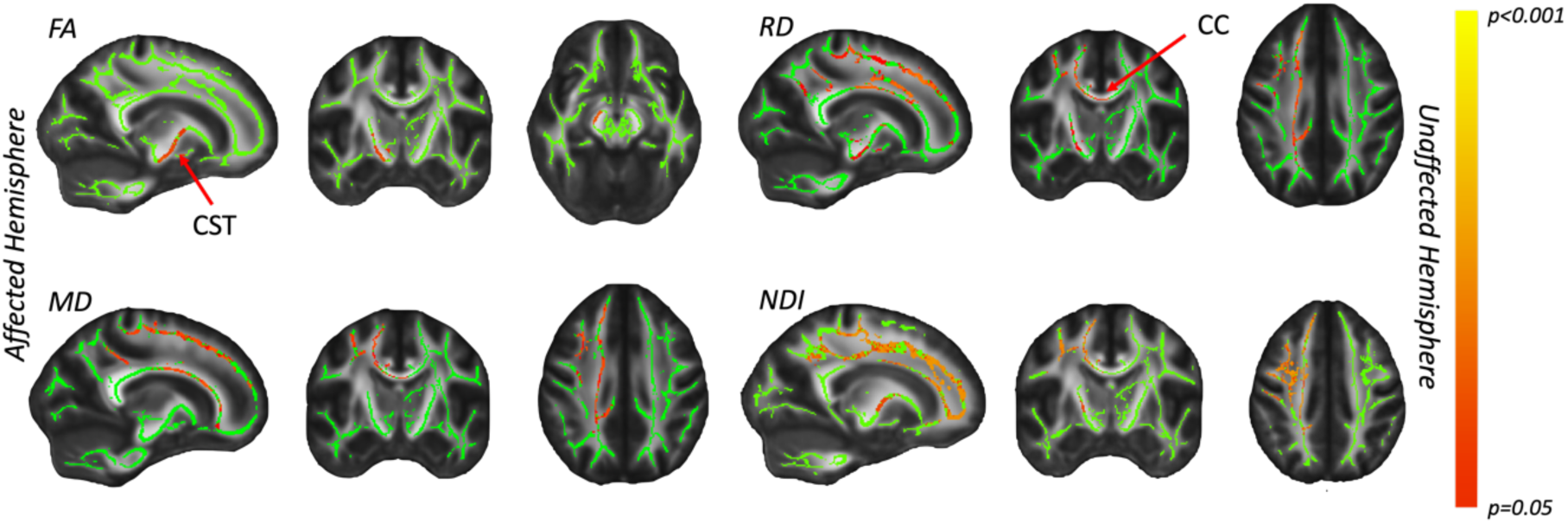
Whole white matter unimodal analysis. Unimodal tests revealed a significant positive association in the expected direction (shown in red) between FA, MD, RD and NDI and stroke survivors score on the UE-FM. (Note MD and RD were negated). Significant voxels were found in the stroke affected hemisphere in the CST, corpus callosum, SLF and superior thalamic radiation.

## Discussion

CST integrity assessed using DWI has frequently been related to motor impairment following stroke^3–7^, and has been suggested to be a promising biomarker of recovery potential in stroke survivors^9^. However, diffusion signals are sensitive to multiple underlying tissue properties including myelination, axon diameter and density, and fibre orientation^10,11^. This makes it difficult to disentangle what biological substrate may underlie any relationship between DWI and impairment or behaviour. The use of multimodal imaging could help to disambiguate underlying biological mechanisms by including imaging modalities with varying sensitivity to different biological tissue properties^19^ in order to identify trends across modalities. In the current study, we used multimodal imaging to further explore relationships between white matter microstructure and motor impairment following stroke, and to shed light on underlying biological mechanisms.

To compare with previous work^3–6^, we first used an ROI approach focused on asymmetry of CST integrity. Using an ROI approach has the advantage of accommodating anatomical variation across individuals, and has previously shown a strong relationship between CST integrity and motor impairment and recovery after stroke^7^. The multimodal NPC analysis using Fisher’s combining function indicated a significant CST asymmetry association with UE-FM, reflecting a joint contribution across all the modalities for this relationship. In line with this finding, the unimodal tests also revealed significant relationships between MD, RD, MTsat and FISO, and UE-FM in the predicted directions (after multiple testing correction across modalities).

MD and NODDI-derived FISO capture unhindered or unrestricted water diffusion and are sensitive to the presence of corticospinal fluid or edema^20,41,42^. MTsat and RD have been proposed to be sensitive to myelination. This assertion has recently been supported by a meta-analysis looking across studies which validated microstructural imaging with myelin histology^26^. This suggests that the previously reported relationship between CST integrity measured with DWI and motor impairment, and the multimodal relationship found in the current paper, may be driven by a combination of a) changes in ‘free water’ content in the CSTs as indexed by MD and FISO, and b) decreases in myelination. This could be interpreted as stroke leading to myelin damage in the stroke affected CST, which therefore leads to fewer barriers to diffusion and in turn increases the amount of unhindered diffusion. Alternatively, the modalities that survived multiple testing correction could also be more sensitive than the other modalities used in the current study^43^ or may be more biologically specific.

In addition to the ROI asymmetry analysis, we carried out a voxelwise asymmetry analysis to examine relationships between asymmetries across the whole white matter and UE-FM. In line with the ROI analysis discussed above, voxels in the CST were again identified in MD, RD and MTsat, as well as in R1. Beyond the CST, the following modalities, MD, RD, MTsat, R1 and NDI, revealed that other brain regions were also found to be related to motor impairment, in particular parts of the SLF, FAT and superior and anterior thalamic radiation (**Figure 2**). These findings are in line with previous work implicating these areas in motor function post stroke^44–46^ and in motor initiation, performance and control^47–49^. In addition, a significant positive association was found between AD and UE-FM in parts of the SLF. AD measures the magnitude of diffusion along the principal eigenvector. Studies in experimental rodent models suggest that reduced AD is associated with axonal damage^22,23^. However, evidence from human studies is more mixed, with several studies reporting increased AD (as well as increased RD) associated with ageing and neurodegeneration^34,35,50^. AD has been found to be particularly difficult to interpret in cases where demyelination, axonal injury and loss, and inflammation occur together. In particular, both RD and AD appear to be increased in chronic conditions with extensive axonal loss^22^, as is the case of stroke. This may explain the common association between increased AD and RD and worse motor impairment found here. However, when correcting across modalities, only greater MTsat asymmetry was associated with worse impairment. This result was found in the SLF, FAT and superior and anterior thalamic radiation and was stronger in these tracts than in the CST (after FWER correction) (see Figure 3). This is unexpected as MTsat is mostly sensitive to myelin and not to fibre organisation or density, like diffusion metrics. These relationships found across different modalities, and particularly MTsat, highlight the potential of using multimodal imaging to uncover further relationships with impairment beyond the CST.

Similarly, in the whole brain white matter voxelwise analysis, greater FA, RD and NDI in the CST in the affected hemisphere was found to be associated with less motor impairment. The same relationship was found for MD, RD and NDI in the corpus callosum, SLF and thalamic radiations in the affected hemisphere. However, none of these metrics survived multiple modalities correction.

Therefore, we were able to replicate previous findings relating CST integrity and motor impairment using both an ROI as well as a whole brain approach. Using multiple modalities, we found that the strongest relationship (surviving multiple testing correction) was found between modalities that measure free water and myelination. This reinforces our understanding of the CST’s role in motor impairment following stroke and sheds some light on the underlying biological mechanisms.

Additionally, we were able to detect relationships in multiple other brain regions. For example, parts of the SLF were highlighted in multiple modalities. Limited time can make it difficult to collect multiple modalities when scanning patients. However, the use of multimodal data allowed us to detect additional relationships that may not have been possible using just one modality alone.

Although FA has been widely used to detect relationships between white matter structure and stroke impairment, in this sample FA seems to be less sensitive overall than other metrics such as MD, RD, MTsat, and FISO in the CST (see Table 1) and when employing voxelwise analysis, FA seems to be mostly sensitive to effects in the CST but not in other tracts (Supplementary Figure 3, Figure 4).

## Limitations

There were limitations in the current study. Firstly, we used an opportunistic sample which was relatively small and heterogeneous. Therefore, caution should be used when interpreting the results and replication with a larger sample is necessary to provide further support to the findings. Additionally, the cross-sectional nature of the data used makes it difficult to determine what the cause behind relationships between MRI modalities and functional impairment are. For example, it is not possible to determine whether these relationships are due to secondary degeneration remote from the lesion site, due to adaptive plasticity or due to pre-existing anatomical differences between individuals. Longitudinal studies are necessary to disentangle these variables.

## Conclusions

In the current study we found multimodal associations when examining CST asymmetry relating our MRI modalities to motor impairment. Modalities sensitive to myelin and free water had the strongest relationship with motor impairment, suggesting that tissue changes involving these biological substrates may underlie the relationships observed between diffusion signals and motor impairment. In addition to the CST, we also found relationships with motor impairment in multiple other brain regions when analysing whole white matter across different modalities. Here, the myelin sensitive metric MTsat, had the strongest relationship with impairment in the SLF, FAT and Superior and anterior thalamic radiation.

The current study highlights the benefits of multimodal data analysis to help shed light on biological mechanisms underlying relationships between brain structure and motor function, as well as the potential to detect a wider range of such relationships.

## Data Availability

Fully anonymized data from this study can be made available on request.

## Acknowledgements

Z.B.S was supported by a Clarendon Scholarship and the Nuffield Department of Clinical Neurosciences. This work was supported by John Fell Award to C.S.B. E.M. was supported by a Wellcome Biomedical Vacation Scholarship. H.J-B was supported by Wellcome Trust grant 110027/Z/15/Z. The Wellcome Centre for Integrative Neuroimaging is supported by core funding from the Wellcome Trust 203139/Z/16/Z.

The funders had no role in the study design, data collection or analysis. The authors would like to thank the radiographers and IT support team at the Wellcome Centre for Integrative Neuroimaging. Fully anonymized data from this study can be made available on request.

<u>For the purpose of open access, the authors have applied a Creative Commons Attribution (CC BY) licence to any Author Accepted Manuscript version arising from this submission.</u>

## References

1. Katan, M. & Luft, A. Global Burden of Stroke. Semin. Neurol. 38, 208–211 (2018).

2. Franceschini, M., La Porta, F., Agosti, M. & Massucci, M. Is health-related-quality of life of stroke patients influenced by neurological impairments at one year after stroke? Eur. J. Phys. Rehabil. Med. 46, 389–399 (2010).

3. Lindenberg, R. et al. Structural integrity of corticospinal motor fibers predicts motor impairment in chronic stroke. Neurology 74, 280–287 (2010).

4. Puig, J. et al. Decreased corticospinal tract fractional anisotropy predicts long-term motor outcome after stroke. Stroke 44, 2016–2018 (2013).

5. Groisser, B. N., Copen, W. A., Singhal, A. B., Hirai, K. K. & Schaechter, J. D. Corticospinal tract diffusion abnormalities early after stroke predict motor outcome. Neurorehabil. Neural Repair 28, 751–760 (2014).

6. Zhu, L. L., Lindenberg, R., Alexander, M. P. & Schlaug, G. Lesion load of the corticospinal tract predicts motor impairment in chronic stroke. Stroke 41, 910– 915 (2010).

7. Stinear, C. M. et al. Functional potential in chronic stroke patients depends on corticospinal tract integrity. Brain 130, 170–180 (2007).

8. Qiu, M. et al. White matter integrity is a stronger predictor of motor function than BOLD response in patients with stroke. Neurorehabil. Neural Repair 25, 275–284 (2011).

9. Boyd, L. A. et al. Biomarkers of Stroke Recovery: Consensus-Based Core Recommendations from the Stroke Recovery and Rehabilitation Roundtable*. Neurorehabil. Neural Repair 31, 864–876 (2017).

10. Zatorre, R. J., Fields, R. D. & Johansen-Berg, H. Plasticity in gray and white: Neuroimaging changes in brain structure during learning. Nat. Neurosci. 15, 528–536 (2012).

11. Sampaio-Baptista, C. & Johansen-Berg, H. White Matter Plasticity in the Adult Brain. Neuron 96, 1239–1251 (2017).

12. Lakhani, B., Hayward, K. S. & Boyd, L. A. Hemispheric asymmetry in myelin after stroke is related to motor impairment and function. NeuroImage Clin. 14, 344–353 (2017).

13. Borich, M. R., MacKay, A. L., Vavasour, I. M., Rauscher, A. & Boyd, L. A. Evaluation of white matter myelin water fraction in chronic stroke. NeuroImage Clin. 2, 569–580 (2013).

14. Cheng, Y. J. et al. Prolonged myelin deficits contribute to neuron loss and functional impairments after ischaemic stroke. Brain 147, 1294–1311 (2024).

15. Okabe, N., Narita, K. & Miyamoto, O. Axonal remodeling in the corticospinal tract after stroke: How does rehabilitative training modulate it? Neural Regen. Res. 12, 185–192 (2017).

16. Liu, Z., Xin, H. & Chopp, M. Axonal remodeling of the corticospinal tract during neurological recovery after stroke. Neural Regen. Res. 16, 939–943 (2021).

17. Linck, P. A. et al. Neurodegeneration of the substantia nigra after ipsilateral infarct: MRI R2∗ mapping and relationship to clinical outcome. Radiology 291, 438–448 (2019).

18. Kuchcinski, G. et al. Thalamic alterations remote to infarct appear as focal iron accumulation and impact clinical outcome. Brain 140, 1932–1946 (2017).

19. Lazari, A. et al. Reassessing associations between white matter and behaviour with multimodal microstructural imaging. Cortex 145, 187–200 (2021).

20. Zhang, H., Schneider, T., Wheeler-Kingshott, C. A. & Alexander, D. C. NODDI: Practical in vivo neurite orientation dispersion and density imaging of the human brain. Neuroimage 61, 1000–1016 (2012).

21. Tae, W. S., Ham, B. J., Pyun, S. B., Kang, S. H. & Kim, B. J. Current clinical applications of diffusion-tensor imaging in neurological disorders. J. Clin. Neurol. 14, 129–140 (2018).

22. Winklewski, P. J. et al. Understanding the physiopathology behind axial and radial diffusivity changes-what do we Know? Front. Neurol. 9, (2018).

23. Song, S. K. et al. Diffusion tensor imaging detects and differentiates axon and myelin degeneration in mouse optic nerve after retinal ischemia. Neuroimage 20, 1714–1722 (2003).

24. Weiskopf, N. et al. Quantitative multi-parameter mapping of R1, PD*, MT, and R2* at 3T: A multi-center validation. Front. Neurosci. 7, 1–11 (2013).

25. Tabelow, K. et al. hMRI – A toolbox for quantitative MRI in neuroscience and clinical research. Neuroimage 194, 191–210 (2019).

26. Lazari, A. & Lipp, I. Can MRI measure myelin? Systematic review, qualitative assessment, and meta-analysis of studies validating microstructural imaging with myelin histology. Neuroimage 230, 117744 (2021).

27. Sanders, Z. B. et al. Self-modulation of motor cortex activity after stroke: A randomized controlled trial. Brain 145, 3391–3404 (2022).

28. Papp, D., Callaghan, M. F., Meyer, H., Buckley, C. & Weiskopf, N. Correction of Inter-Scan Motion Artifacts in Quantitative R1 Mapping by Accounting for Receive Coil Sensitivity Effects. 1485, 1478–1485 (2016).

29. Tariq, M., Schneider, T., Alexander, D. C., Gandini Wheeler-Kingshott, C. A. & Zhang, H. Bingham-NODDI: Mapping anisotropic orientation dispersion of neurites using diffusion MRI. Neuroimage 133, 207–223 (2016).

30. Castella, R. et al. Controlling motion artefact levels in MR images by suspending data acquisition during periods of head motion. Magn. Reson. Med. 80, 2415–2426 (2018).

31. Avants, B., Tustison, N. & Song, G. Advanced Normalization Tools (ANTS). Insight J. 1–35 (2009).

32. Smith, S. M. et al. Tract-based spatial statistics : Voxelwise analysis of multi-subject diffusion data. 31, 1487–1505 (2006).

33. Winkler, A. M., Ridgway, G. R., Webster, M. A., Smith, S. M. & Nichols, T. E. Permutation inference for the general linear model. Neuroimage 92, 381–397 (2014).

34. Korbmacher, M. et al. Brain-wide associations between white matter and age highlight the role of fornix microstructure in brain ageing. Hum. Brain Mapp. 44, 4101–4119 (2023).

35. Sexton, C. E. et al. Accelerated changes in white matter microstructure during aging: A longitudinal diffusion tensor imaging study. J. Neurosci. 34, 15425–15436 (2014).

36. Winkler, A. M. et al. Non-parametric combination and related permutation tests for neuroimaging. Hum. Brain Mapp. 37, 1486–1511 (2016).

37. Winkler, A. M., Ridgway, G. R., Douaud, G., Nichols, T. E. & Smith, S. M. Faster permutation inference in brain imaging. Neuroimage 141, 502–516 (2016).

38. Lebel, C. et al. NeuroImage Diffusion tensor imaging of white matter tract evolution over the lifespan. Neuroimage 60, 340–352 (2012).

39. Bennett, I. J. & Madden, D. J. Disconnected aging: Cerebral white matter integrity and age-related differences in cognition. Neuroscience 276, 187–205 (2014).

40. Callaghan, M. F. et al. Widespread age-related differences in the human brain microstructure revealed by quantitative magnetic resonance imaging. Neurobiol. Aging 35, 1862–1872 (2014).

41. Palacios, E. M. et al. The evolution of white matter microstructural changes after mild traumatic brain injury: A longitudinal DTI and NODDI study. Sci. Adv. 6, 1–11 (2020).

42. Heath, F., Hurley, S. A., Johansen-Berg, H. & Sampaio-Baptista, C. Advances in noninvasive myelin imaging. Dev. Neurobiol. 78, 136–151 (2018).

43. Wenger, E. et al. Reliability of quantitative multiparameter maps is high for magnetization transfer and proton density but attenuated for R1 and R2* in healthy young adults. Hum. Brain Mapp. 43, 3585–3603 (2022).

44. Schulz, R. et al. Parietofrontal motor pathways and their association with motor function after stroke. Brain 138, 1949–1960 (2015).

45. Jacquemont, T. et al. Association between superior longitudinal fasciculus, motor recovery, and motor outcome after stroke: a cohort study. Front. Neurol. 14, (2023).

46. Schaechter, J. D. et al. Microstructural Status of Ipsilesional and Contralesional Corticospinal Tract Correlates with Motor Skill in Chronic Stroke Patients. Hum. Brain Mapp. 30, 3461–3474 (2009).

47. Budisavljevic, S. et al. The role of the frontal aslant tract and premotor connections in visually guided hand movements. Neuroimage 146, 419–428 (2017).

48. Kinoshita, M. et al. Role of fronto-striatal tract and frontal aslant tract in movement and speech: an axonal mapping study. Brain Struct. Funct. 220, 3399–3412 (2015).

49. Rodríguez-Herreros, B. et al. Microstructure of the superior longitudinal fasciculus predicts stimulation-induced interference with on-line motor control. Neuroimage 120, 254–265 (2015).

50. Barrick, T. R., Charlton, R. A., Clark, C. A. & Markus, H. S. White matter structural decline in normal ageing: A prospective longitudinal study using tract-based spatial statistics. Neuroimage 51, 565–577 (2010).

