## supplementary materials for "Quantitative multimodal microstructural imaging associations with chronic stroke motor impairment"

**Supplementary Material**

**Supplementary Table 1*:*** *corrected and uncorrected t-stat and p-values for each modality in the ROI-based CST asymmetry analysis for the opposite contrast.*

|  | **t-stat** | **uncorrected p-value** | **corrected p-value** |
| --- | --- | --- | --- |
| **FA** | -2.6872 | 0.9920 | 1.000 |
| **MD** | -2.8445 | 0.9934 | 1.000 |
| **AD** | 1.8454 | 0.0414 | 0.2220 |
| **RD** | -3.2616 | 0.9982 | 1.000 |
| **MTsat** | -3.0346 | 0.9954 | 1.000 |
| **R1** | -2.6417 | 0.9886 | 1.000 |
| **R2*** | 0.3155 | 0.3816 | 0.9292 |
| **NDI** | -2.7493 | 0.9930 | 1.000 |
| **ODI** | -1.7056 | 0.9452 | 1.000 |
| **fiso** | -2.9087 | 0.9952 | 1.000 |

*Results without covariates*

PALM was used to carry out a voxelwise multi- and uni-modal analysis of relationships between UE-FM scores and white matter asymmetry. When testing for a joint effect of the ten modalities using Fisher’s combining function, a significant effect was found in the CST and the superior thalamic radiation (**Supplementary Figure 2**), with greater asymmetry in these areas being associated with a lower score on the UE-FM. Significant effects were also detected on the unimodal tests; a negative correlation was found with FA, MD, RD, MTsat, R1, R2* and NDI, and a positive correlation with AD (against the hypothesis) (**Supplementary Figure 4**). MD, RD, MTsat, R1 and NDI correlations survive multiple comparison correction across modalities (**Supplementary Figure 5**).

When testing multimodal relationships between the whole brain white matter and UE-FM, there was a significant result in parts of the stroke affected SLF (see **Supplementary Figure 6**). Additionally, significant unimodal results were found in FA, MD, AD (inverse contrast), RD and NDI (see **Supplementary Figure 7**). None of these results survived correction across modalities.

*Supplementary Figures*

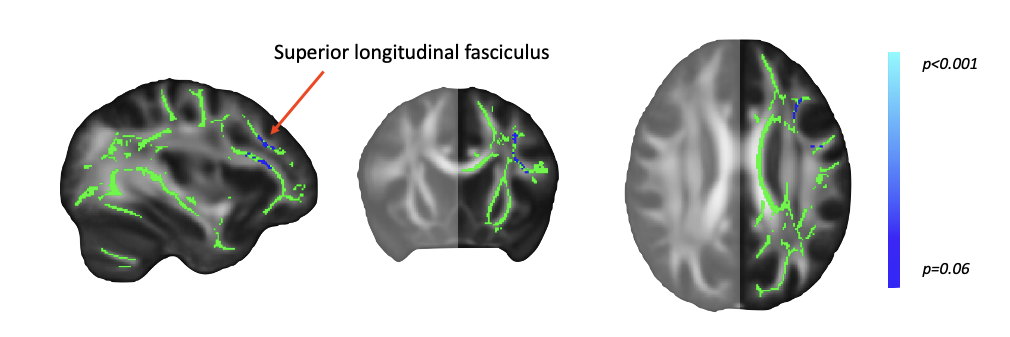

**Supplementary Figure 1:** Multimodal voxelwise asymmetry analysis with covariates. The test failed to reach significance when testing for a negative correlation between white matter asymmetry across the modalities and stroke survivors score on the UE-FM (peak voxel p=0.057). Peak voxels were found in the areas of the superior longitudinal fasciculus (SLF).

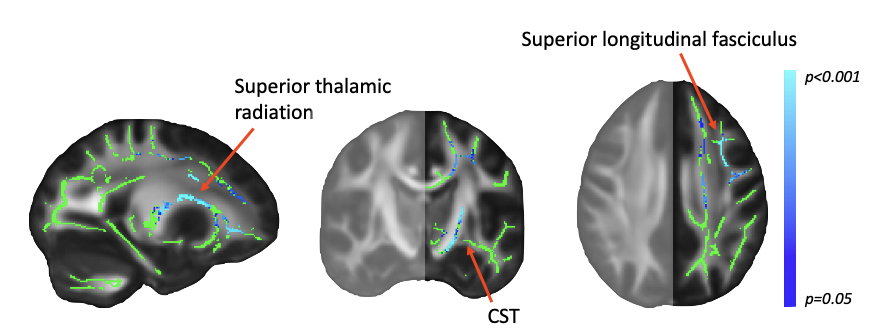

**Supplementary Figure 2:** Multimodal voxelwise asymmetry analysis excluding covariates. When no covariates were included in the model, the test revealed a significant negative correlation between white matter asymmetry and stroke survivors UE-FM score (p<0.05 corrected). Significant voxels were located in the Cortical Spinal Tract (CST), SLF and Superior thalamic radiation. Significant clusters are only projected in one hemisphere as asymmetry analysis considers the difference between affected hemisphere minus the unaffected.

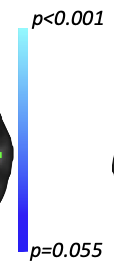

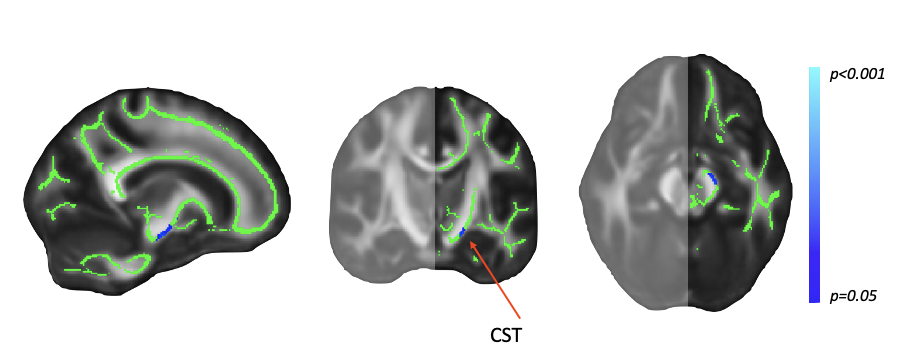

**Supplementary Figure 3:** Voxelwise asymmetry unimodal analysis with covariates. A trend was found towards a negative correlation between FA asymmetry and score on the UE-FM in the CST (peak voxel p=0.051). Significant clusters are only projected in one hemisphere as asymmetry analysis considers the difference between affected hemisphere minus the unaffected.

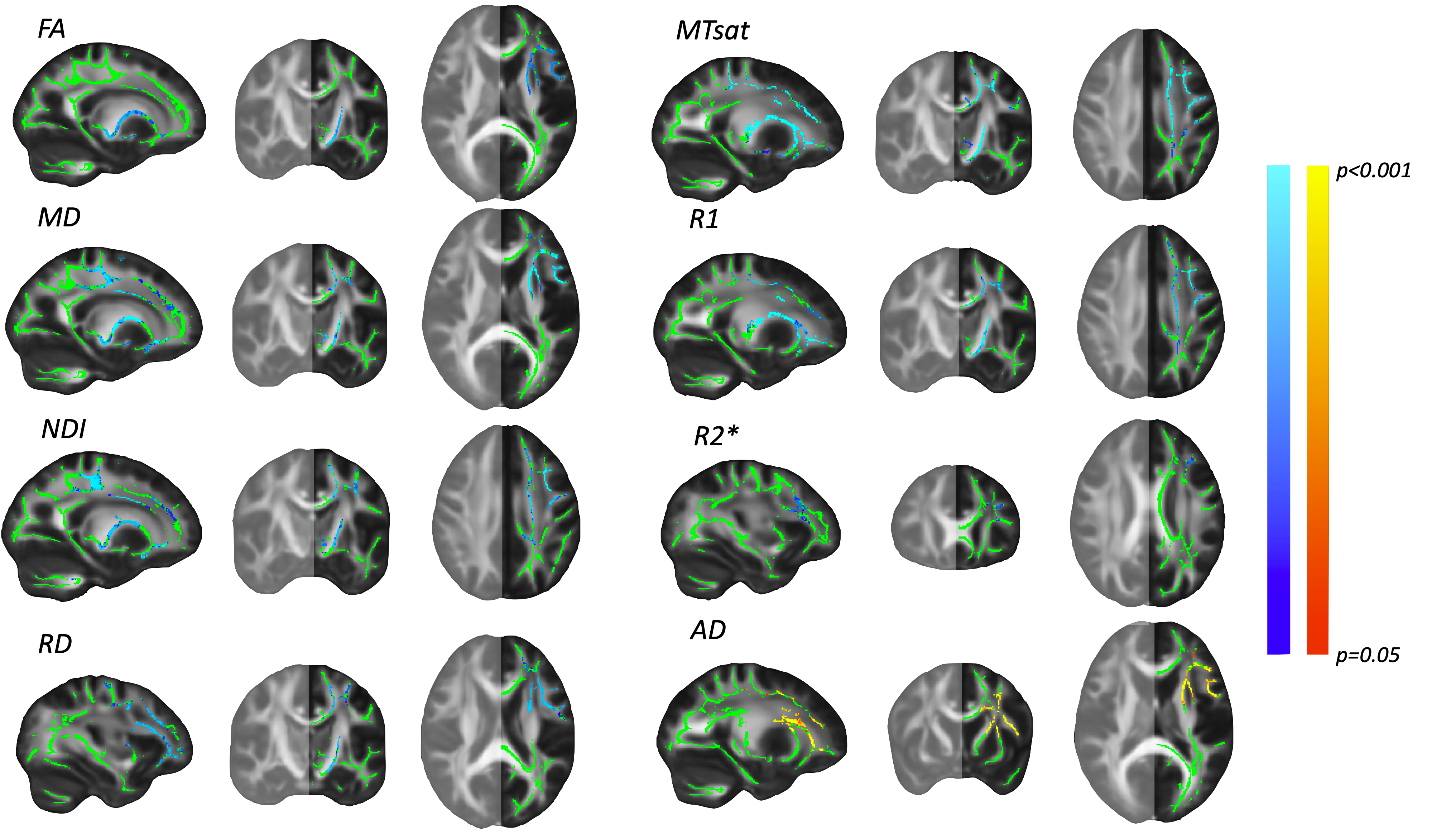

**Supplementary Figure 4**: Voxelwise asymmetry unimodal analysis excluding covariates. Significant negative correlations (family-wise error corrected; shown in blue) were found in the expected direction between FA, MD, RD, MTsat, R1, R2* and NDI asymmetry, and the UE-FM score (note *RD and MD values were negated).* Significant voxels were found in several motor related areas including the CST, SLF, FAT and superior and anterior thalamic radiation. Additionally, a significant positive correlation (shown in red) was found between AD and the UE-FM score. Significant clusters are only projected in one hemisphere as asymmetry analysis considers the difference between affected hemisphere minus the unaffected.

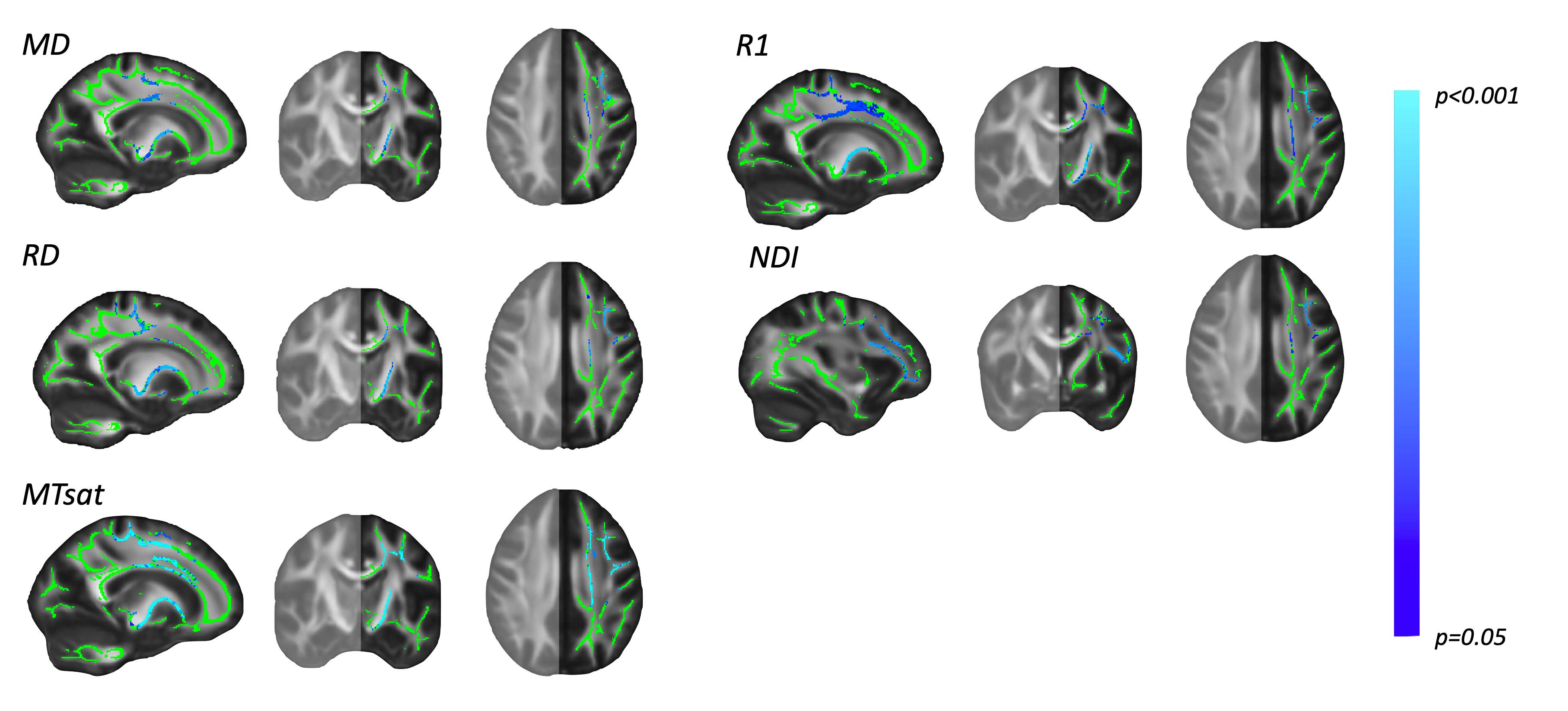

**Supplementary Figure 5**: Voxelwise asymmetry unimodal analysis excluding covariates, after FWER-correction across modalities. Significant negative correlations were found in the expected direction (shown in blue) between MD, RD, MTsat, R1 and NDI asymmetry, and the UE-FM score following multiple comparison correction across modalities (note *RD and MD values were negated).* Significant voxels were found in several motor related areas including the CST, SLF, and superior and anterior thalamic radiation. Significant clusters are only projected in one hemisphere as asymmetry analysis considers the difference between affected hemisphere minus the unaffected.

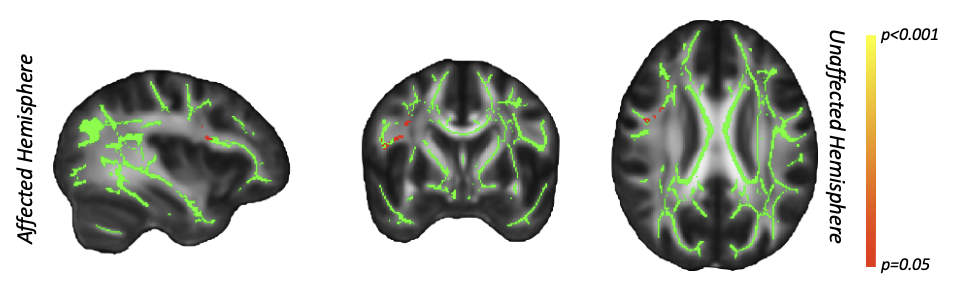

**Supplementary Figure 6**: Multimodal whole white matter analysis without covariates. The test revealed significant positive correlation (shown in red) on the across the MRI modalities and stroke survivors UE-FM score. Significant voxels were located in the stroke affected hemisphere in the SLF.
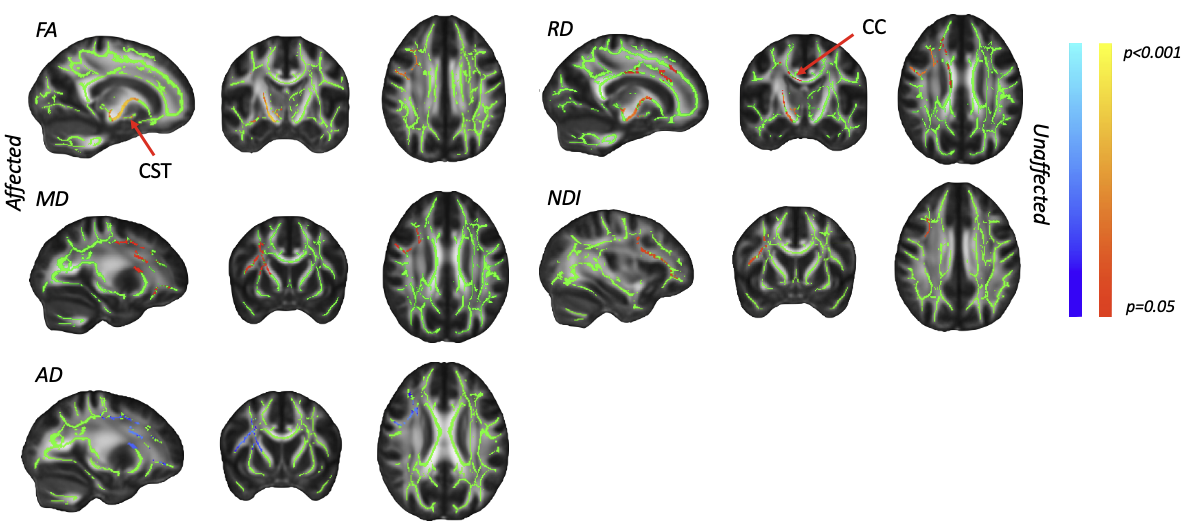

**Supplementary Figure 7**: Whole white matter voxelwise unimodal analysis without covariates. Significant positive correlations (family-wise error corrected; shown in red) were found in the expected direction (between FA, MD, RD, and NDI, and UE-FM score (note *RD and MD were negated).* Significant voxels were found in several motor related areas including the CST, CC, SLF and FAT. Additionally, against the hypothesis a significant negative correlation (shown in blue) was found between AD and the UE-FM. None of the results survived multiple comparison correction across modalities.
